# Highly Effective Inactivation of SARS-CoV-2 by Conjugated Polymers and Oligomers

**DOI:** 10.1101/2020.09.29.20204164

**Authors:** Florencia A. Monge, Pradeepkumar Jagadesan, Virginie Bondu, Patrick L. Donabedian, Linnea Ista, Eva Y. Chi, Kirk S. Schanze, David G Whitten, Alison M. Kell

**Affiliations:** Center for Biomedical Engineering, University of New Mexico; Biomedical Engineering Graduate Program, University of New Mexico; Department of Chemistry; University of Texas at San Antonio; Nanoscience and Microsystems Engineering Graduate Program, University of New Mexico; Department of Chemical and Biological Engineering, University of New Mexico; Department of Chemistry and Chemical Biology, University of New Mexico; Department of Molecular Genetics and Microbiology, University of New Mexico School of Medicine

## Abstract

The current Covid-19 Pandemic caused by the highly contagious SARS-CoV-2 virus has proven extremely difficult to prevent or control. Currently there are few treatment options and very few long-lasting disinfectants available to prevent the spread. While masks and protective clothing and “social distancing” may offer some protection, their use has not always halted or slowed the spread. Several vaccines are currently undergoing testing; however there is still a critical need to provide new methods for inactivating the virus before it can spread and infect humans. In the present study we examined the inactivation of SARS-CoV-2 by synthetic conjugated polymers and oligomers developed in our laboratories as antimicrobials for bacteria, fungi and non-enveloped viruses. Our results show that we can obtain highly effective light induced inactivation with several of these oligomers and polymers including irradiation with near-UV and visible light. With both the oligomers and polymers, we can reach several logs of inactivation with relatively short irradiation times. Our results suggest several applications involving the incorporation of these materials in wipes, sprays, masks and clothing and other Personal Protection Equipment (PPE) that can be useful in preventing infections and the spreading of this deadly virus and future outbreaks from similar viruses.

## INTRODUCTION

It is hard to overstate the devastation brought to the world by the SARS-CoV-2 virus through the highly contagious Covid-19 disease. In contrast, of the six previously encountered human coronaviruses, four are common and remain active but pose only minor threats to world health.^1^ Three recent (since 2000) coronaviruses originally infected animals but have jumped to humans and pose existential threats to human health. SARS coronavirus (SARS-CoV) emerged in 2002 and caused Severe Acute Respiratory Syndrome (SARS). SARS-CoV emerged in an outbreak in February 2003 in Asia and rapidly spread to two dozen countries in Asia, South America, North America and Europe before being contained.^2-7^ According to the World Health Organization (WHO), there were 8,098 cases worldwide and of those 774 died.^8^ In the US there were 8 verified cases and no deaths and there have been no cases of SARS subsequent to 2003. Middle East Respiratory Syndrome (MERS) emerged in Saudi Arabia in 2012 and has since spread to several countries, including the US. MERS causes severe respiratory symptoms, including death. There have been two nonfatal confirmed cases in the US in May 2014. MERS is a threat to travelers to the Middle East and travel advisories are posted on the CDC Website. In contrast to the SARS and MERS, Covid-19 has spread to all inhabited parts of the world, excluding Antarctica. As of September 5, 2020, there are 26,468,000 cases and 871,000 deaths worldwide, and of those 6,095,007 cases and 185,687 deaths have occurred in the US.^8^

The major attempt at controlling the spread of Covid-19 includes several efforts to produce vaccines. Although promising candidates are already in human clinical trial, it is likely that unforeseen problems may delay the availability of a safe and effective vaccine and its widespread employment. Several diagnostic tests for of SARS-CoV-2 have been developed; however, uneven test distribution and delays in obtaining results have hindered our knowledge of viral spread and hampered out ability to develop an effective response. Several conventional disinfectants are active against the SARS-CoV-2 virus and the most commonly used of these are bleach, hydrogen peroxide, and alcohol solutions. While these disinfectants are effective for use as cleaning solutions and wipes, the volatility and/or corrosivity of the “active agents” limit prolonged sterilization of surfaces or objects by these reagents.

Here we present an alternative approach to long lasting disinfection: cationic phenylene ethynylene polymers and oligomers (conjugated electrolytes).^9-15^ Over the past decade we and others have demonstrated the effectiveness of these compounds and derivative materials against bacteria, fungi, biofilms, and viruses.^16-26^ The antimicrobial activity of these compounds includes both light-activated and dark-active pathways. Encouragingly, these materials exhibit relatively low toxicity against mammalian cells or the human skin.^27, 28^ Depending upon the selection of materials and properties such as solubility and stability, it is possible to control the lifetime of these materials in coatings, with photodegradation as the main route of inactivation.^29^ The most common byproducts of photodegradation, due to oxidative cleavage, are aldehydes and carboxylic acids that are unlikely to damage the environment.^29^ In addition, these materials can also bind selectively with certain proteins or protein fragments and with light-irradiation, the conjugated electrolytes may photosensitize the oxidation of proteins at the specific binding sites.^30^

The present paper reports a pilot study of five phenylene ethynylene materials and compounds as inactivators of the SARS-CoV-2 virus in aqueous suspensions. The five materials were chosen as representatives of a much larger group of polymeric and oligomeric conjugated electrolytes that have been previously shown to be highly active against microbes.^10, 11, 16, 18, 31-33^ In this study, samples of each material in solution was incubated with an aqueous suspension of SARS-CoV-2. The samples were incubated in the dark and under UV-visible light irradiation in a photoreactor. After incubation for increasing amounts of time, the samples were analyzed for virus activity. As detailed below, it was found that all five of the tested materials were effective against the SARS-CoV-2 virus, but with effectiveness that differed significantly among the tested group.

## RESULTS

### Oligomer and Polymer Structures and Rationale for Selection

The structures of the five materials tested are shown in Figure 1. Three are oligomeric phenylene ethynylenes (OPEs, compounds **1, 2**, and **3**) and two are cationic conjugated polymers (**poly-4** and **poly-5**).

**Figure 1.**
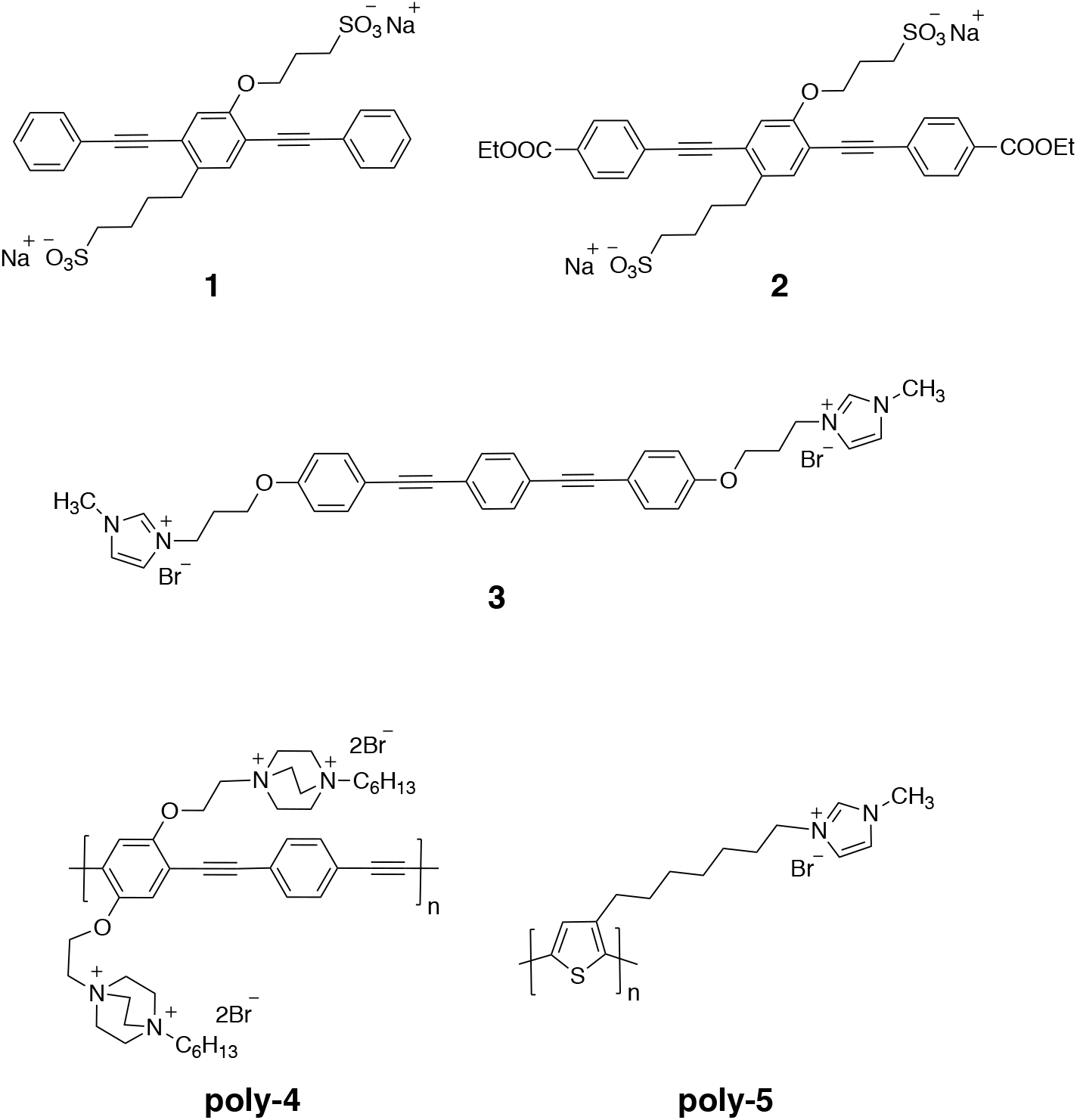
Structures of oligomers (**1**-**3**) and polymers (**poly-4**, and **poly-5**) used in this study.

The synthesis of **3** is reported in Supplemental Material and the syntheses of all other materials have been reported in the literature.^31, 34-36^ As shown in Figure 2, all of the materials absorb in the near-UV and/or visible regions of the spectrum. The final concentration of materials chosen for this study was 10 µg/mL in solutions that contain an active compound and SARS-CoV-2. This concentration was selected to give moderate absorption throughout the sample and yet minimizing a potential “inner filter” effect.^37, 38^ The fact that the **poly-4** absorbs much more strongly at this concentration suggests the possibility that an “inner filter” effect could diminish its light-activated viral inactivation activity.

**Figure 2.**
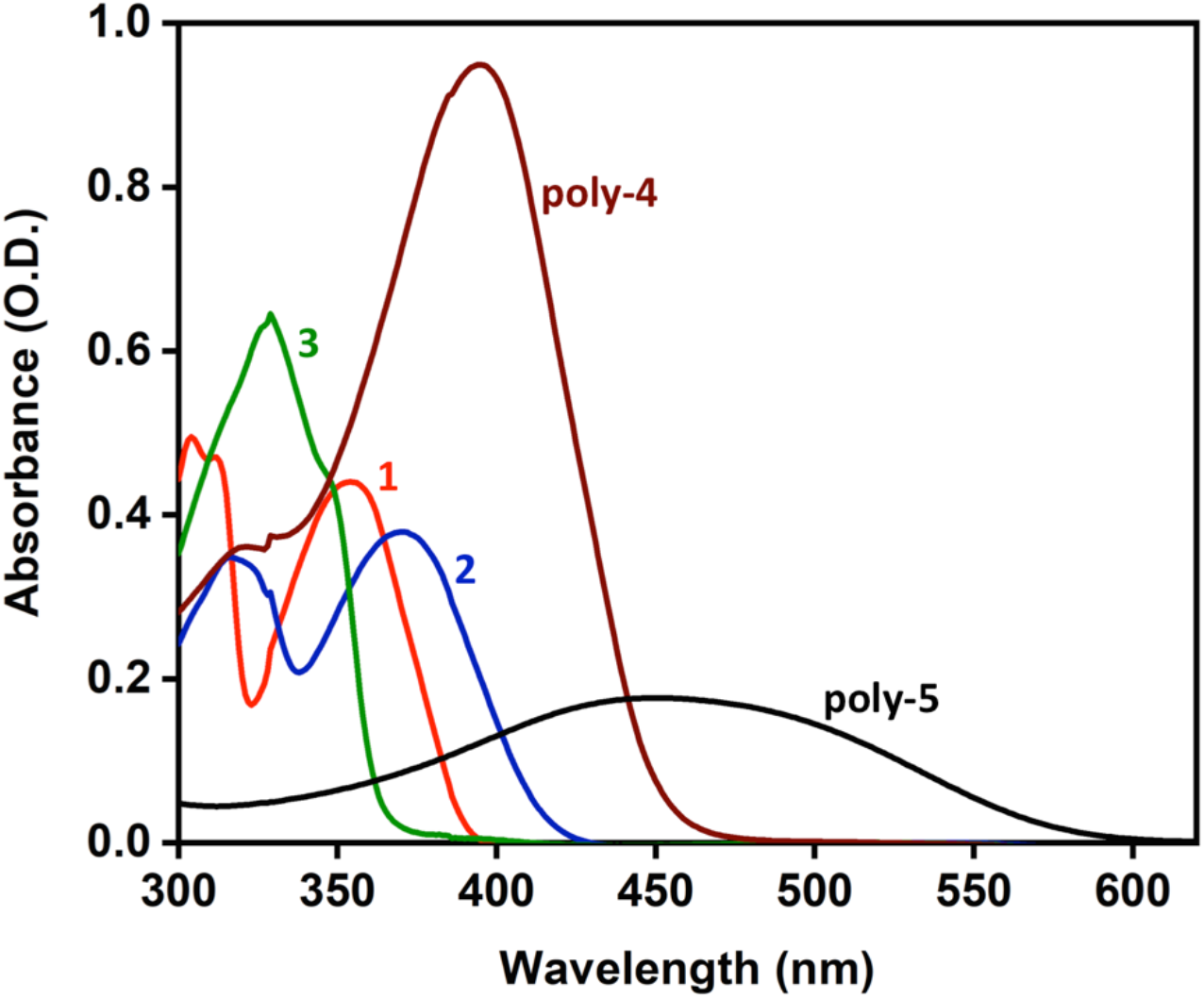
Absorption spectra of the five phenylene ethynylene materials at 10 µg/mL in water. Spectra were recorded on a Lambda 35 U/Vis spectrometer (PerkinElmer, Waltham, MA) in quartz cuvettes (PerkinElmer, Waltham, MA).

Our previous studies testing the antiviral activities of oligomeric and polymeric phenylene ethynylene materials were performed with the non-enveloped RNA bacteriophages MS2 and T4.^18^ In contrast, SARS-CoV-2 is an enveloped virus containing several proteins that shield its internal RNA. Recent reports suggest that the virus has an acidic coat and might be reactive towards negatively charged reagents.^39^ We hypothesize that the viral envelope might have both positively and negatively charged regions that could interact with charged compounds such as OPEs and PPEs. Thus, the selection of the test compounds for the pilot project was in-part based on charge: oligomers **1** and **2** are negatively charged. Moreover, oligomer **2** has been shown to associate strongly with several proteins^40-42^ and in fact, induces protein oxidation upon irradiation.^30^ The cationic polymer **poly-4** has become our workhorse compound for antimicrobial work.^33^ Cationic oligomer **3** has not been studied extensively but was selected for investigation due to our findings that phenylene ethynylene polymers, such as **poly-5** containing pendant imidazolium groups are unusually reactive in light induced bacterial killing.^20, 32^

### Photoactive Oligomers Potently inhibit SARS-CoV-2

SARS-CoV-2 that causes Covid-19 efficiently infects and causes cytopathic effects in Vero E6 cells.^43^ Vero E6 cells have an impaired type I IFN response and are therefore highly susceptible to virus infection and cell death.^44, 45^ Vero cells stained with crystal violet are commonly used to define infectious particle concentrations as expressed in plaque forming units (pfu) in viral stocks.^46^ A representative sample with a low pfu concentration is shown in Figure 3.

**Figure 3.**
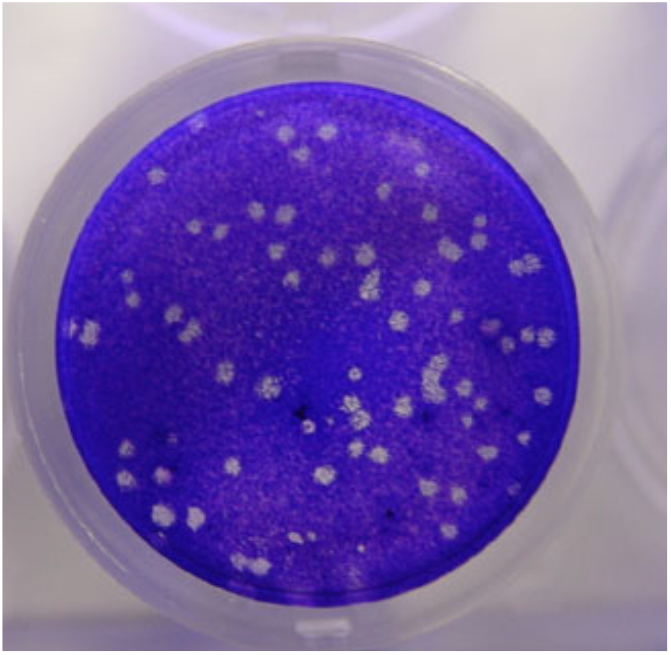
Crystal violet stain of a Vero E6 cell monolayer 3 days post SARS-CoV-2 infection. A viral plaque, which appears as white circular features in the image, begins when a virus infects a cell within the cell monolayer. The virus infected cell subsequently lyses and spreads the infection to adjacent cells where the infection-to-lysis cycle is repeated. The infected cell area creates a plaque, an area of dead cells surrounded by uninfected, live cells, which can be seen by adding a crystal violet solution that colors the cytoplasm of healthy cells.

We first tested the near UV light (300-400 nm) activated inhibitory effect of oligomers **1, 2** and **3** against SARS-CoV-2 infection on Vero cells. Figure 4 shows the reduction in pfu when SARS-CoV-2 is mixed in cell culture media with the oligomers and incubated with near UV light irradiation. Compounds **1** and **3** demonstrated immediate activity and ablated any viral plaque formation as fast as 15-20 minutes of irradiation for oligomer **1** and 10 minutes irradiation for oligomer **3** in these conditions (Figure 4). While not as striking, oligomer **2** also showed a clear ability to decrease plaque formation by 30 minutes incubation and irradiation (Figure 4). Importantly, these oligomers demonstrated no antiviral activity when incubated in the absence of light and demonstrated no cytopathic effects when added to Vero cell monolayers in the absence of virus (data not shown).

**Figure 4.**
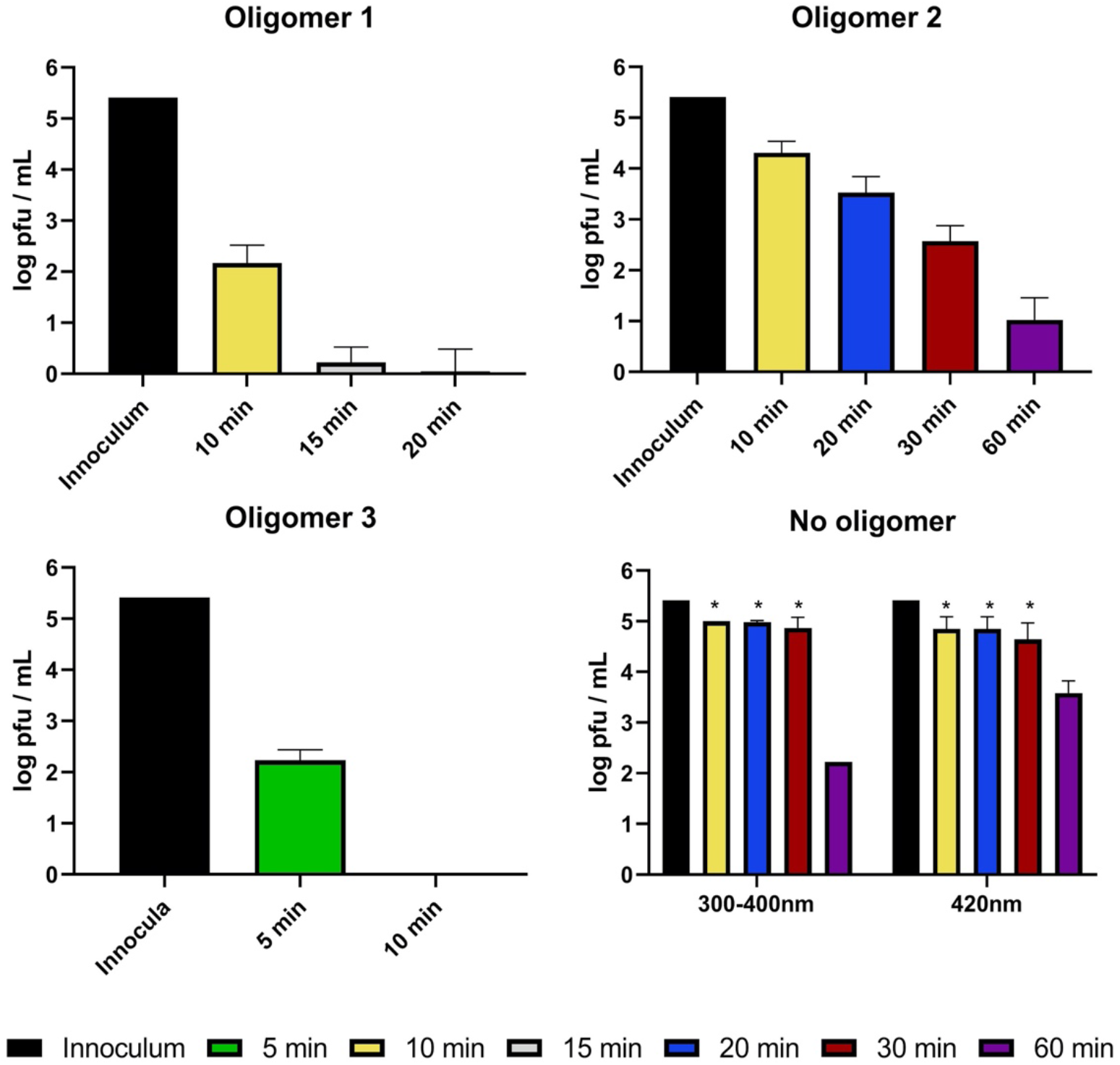
Antiviral activity of photoactive oligomers against SARS-CoV-2 with near UV light irradiation for the indicated time periods. 10 μg/mL of each oligomer was incubated with 2.6×10^5^ pfu/mL SARS-CoV-2 and irradiated. Viral titer was quantified by plaque assay on Vero E6 cells. Asterisks (*) denote that upper limit of detection was reached for plaque assays with these samples. Values shown are the average titer from at least three independent experiments (±SEM).

We then expanded our investigations to include photoactive polymers **poly-4** and **poly-5** which, unlike the oligomers, absorb in the visible region as well as the near-UV (Figure 2), making them likely to be active at these wavelengths as well. As shown in Figure 5A, when incubated with SARS-CoV-2 at a concentration of 10 μg/mL, **poly-**4 reduced plaque formation by almost 1.5 logs after 20 minutes of exposure to near UV light, but this reduction is minimally greater than that of the control with no compound added when exposed to either near UV or visible light. In contrast, **poly-5** is capable of effectively eliminating viral plaque formation in as little as 30 minutes incubation in near UV light and reducing viral titers by nearly 5 logs with 60 minutes of irradiation in the visible light (Figure 5B). Because of the potential “inner filter” effect from **poly-4** and **poly-5** during light incubation, we tested their ability to inhibit SARS-CoV-2 plaque formation at a lower concentration of 5 μg/mL under the same conditions. This lower concentration of either polymer demonstrated minimal differences in viral titer compared to the inhibition observed using 10 μg/mL (Figure 5C). Similar to the oligomer results, we observed neither cytopathic effects of the polymers alone on Vero cells nor reduction in viral titer by plaque assay when polymers were incubated with the virus in the dark (data not shown). We therefore conclude that the “inner filter” effect is not interfering with the antiviral activity of **poy-4** and **poly-5** in these conditions. Further, **poly-5** demonstrated effective inhibition of SARS-CoV-2 in both near UV and visible ranges of light, making it a strong candidate for further study and potential use to combat viral environmental contamination.

**Figure 5.**
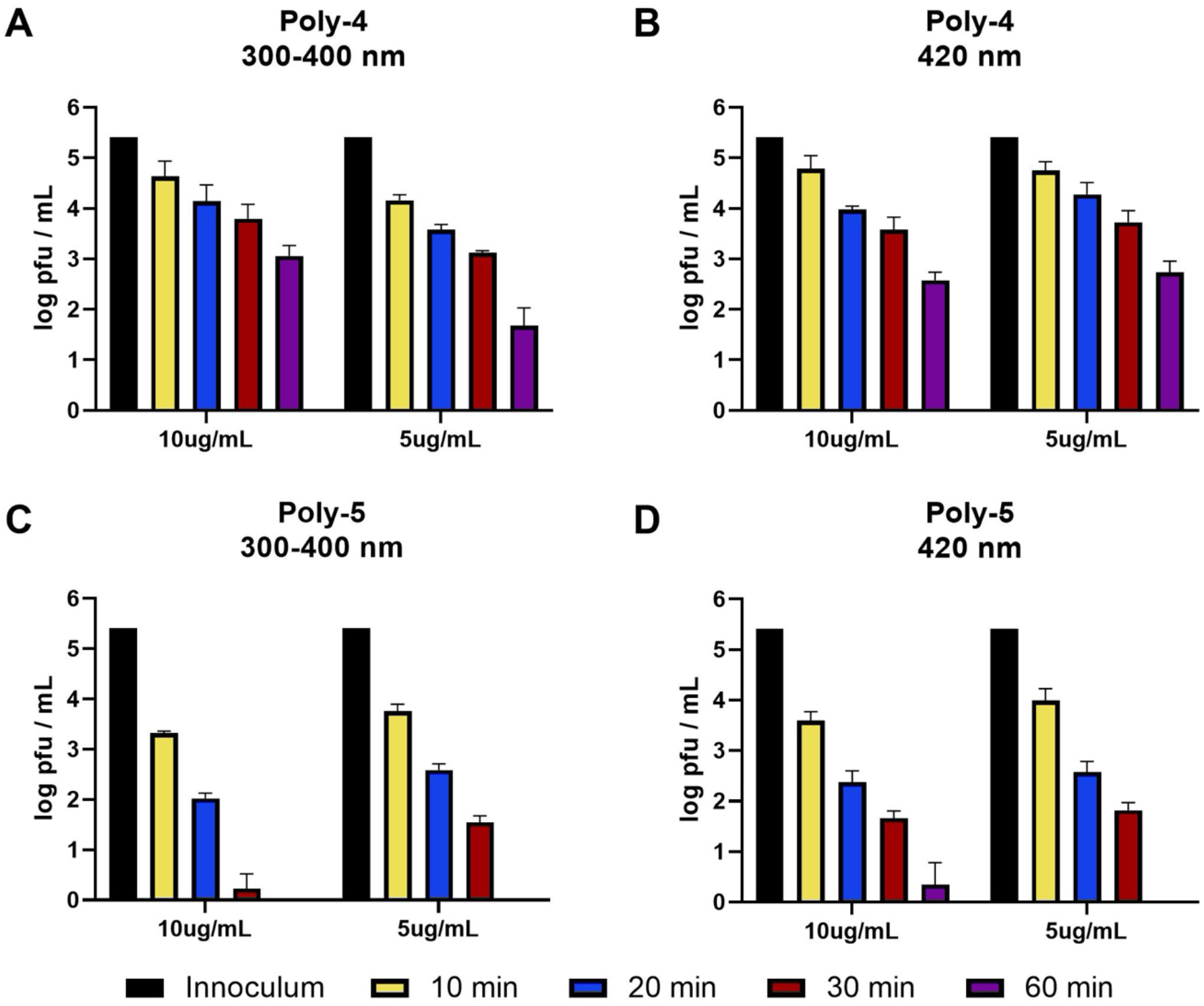
Antiviral activity of photoactive polymers against SARS-CoV-2. **Poly-4** (A, B) or **poly-5** (C, D) was incubated with 2.6×10^5^ pfu/mL SARS-CoV-2 with near UV light or visible light irradiation for the noted times. Viral titer was quantified by plaque assay on Vero E6 cells. Values plotted are the average titer calculated from at least three independent experiments (±SEM).

## DISCUSSION

The most important and significant result of this study is that all five materials tested show antiviral activity against SARS-CoV-2 under irradiation with light absorbed by the specific material. The three oligomers are active when irradiated with near UV light and the two polymers are active under both visible and near-UV irradiation. Both anionic (**1** and **2)** and cationic (**3)** oligomers are active; this is reasonable for non-covalent binding of the oligomers to protein components, likely the viral spike proteins, of the SARS-CoV-2. We have shown in a previous simulation study that the binding of phenylene ethynylene-based oligomers to the MS2 virus capsid protein assembly is mediated by strong van der Waals interactions between the hydrophobic OPE backbone and the capsid protein and strong electrostatic interactions between the OPE charged side chains and charged residues on the capsid surface.^47^ A combination of hydrophobic and electrostatic interactions between the oligomers and polymers and the SARS-CoV-2 spike proteins likely contribute to the compounds’ antiviral activities in this study. Of the 4 structural protein components, the membrane protein M is a likely target for both oligomers and polymers to bind and be in close proximity to interior RNA.^3^ As shown in Figure 4D, when suspensions of SARS-CoV-2 are irradiated with near UV or visible light without oligomers or polymers present, there is some inactivation of the virus, especially upon prolonged irradiation with near UV. However, it is clear that most of the inactivation of the virus under irradiation is due to excitation of the oligomers and polymers.

None of the five materials studied thus far exhibit antiviral activity in the dark. This is not surprising since SARS-CoV-2 is an enveloped virus and in the dark the interactions between oligomers and polymers and membrane proteins are not expected to be strong enough to denature the protein or break covalent bonds. However, it seems to be the case that binding brings the oligomers and polymers sufficiently close to the virus such that the reactive oxygen species (ROS) generated at the binding sites on the virus can penetrate the virus and cause damages to the membrane coat protein M, RNA and/or inner proteins. Quite likely, the smaller oligomers may penetrate into the interior of the virus perhaps even binding to the N-protein that is directly connected to the virus RNA.^3^ Since we have shown in other studies that both negatively and positively charged OPEs can bind with misfolded proteins or protein fragments,^40-42^ binding of the polymers and oligomers used in this study with virial proteins and subsequent generation of singlet oxygen by irradiation close to the RNA seems a likely path for virus inactivation.^18^ The observation that the cationic quaternary ammonium **poly-4** is inactive in the dark is a harbinger that classic quaternary ammonium surfactants may not be particularly active against the virus.

As mentioned previously and detailed in Figure 4, anionic oligomer **1** and cationic oligomer **3** show rapid and effective inactivation of SARS-CoV-2 under irradiation with near UV light with essentially complete inactivation in 10-15 min, while anionic oligomer **2** shows much slower inactivation under the same conditions. While oligomer **2** has been shown in other studies to bind strongly to misfolded proteins and fragments,^40-42^ it seems reasonable as **2** is more hydrophilic than **1** or **3** and better solubilized in water but less able to penetrate into the interior of the virus. The curious effect that oligomer **2** is less active than **1** may also be associated with the fact that the ester groups have been shown to give rise to rapid excited state quenching of the oligomer in water. This quenching may reduce the ability of the compound to sensitize reactive oxygen species.^48, 49^

For the two polymers used in this study, irradiation with either near UV or visible light results in inactivation of the SARS-CoV-2 virus as shown in Figure 5. **Poly-5** shows rapid and efficient inhibition of the virus with essentially complete inactivation in 20-30 min under either near UV or visible irradiation at levels of both 5 and 10 µg/mL. **Poly-4** is significantly less reactive than **Poly-5**. At this point it is premature to speculate on the cause of the differences. An attractive feature of both polymeric systems examined is that they are reactive under visible irradiation and potentially can be “passive” antiviral agents under ambient irradiation conditions.

It is worth considering the practical applications of these materials. We can envision that these materials can be used in the prevention of Covid-19 and other virus-based diseases as well as for future virus threats. In previous studies we have shown that similar materials prepared in our laboratories can be incorporated into surface coatings. For example, textiles where compound are covalently attached via electrospinning or non-covalently incorporated by adsorption have exhibited broad spectrum antimicrobial properties.^9, 50^ Other studies have shown that these materials are not likely to harm the environment from their degradation byproducts and also not harmful to human skin or to several other types of mammalian cells.^27, 28^ It seems likely that these materials, used as pure components or mixtures in wipes, sprays, personal protective equipment (PPE) items, clothing for athletes, warfare fighters, and paints and coatings, can provide lasting disinfection of hard surfaces in rooms, vehicles, outdoor and indoor spaces.

## CONCLUSIONS

In this pilot study, we have tested five representative conjugated oligomers and polymers from an array of phenylene ethynylene-based cationic and anionic conjugated materials against SARS-CoV-2, the virus that causes the Coved-19 disease. All five of the materials investigated show moderate to very strong inactivation of the virus on irradiation with near-UV or visible light. Although the oligomers and polymers are active under irradiation, they do not inactivate the virus in the dark. The antiviral activities of the compounds are likely due to binding of the compounds to viral proteins that brings the compounds in close proximity to the virus, followed by light-activated singlet oxygen and ROS generation that ultimately damage and inactivate the virus. Our findings open the way towards a host of applications that may mitigate disease infections from SARS-CoV-2 as well as current and future virus threats to human health.

## MATERIAS AND METHODS

### Cells and viruses

Vero E6 cells (ATCC, CRL-1586) were cultured in Dulbecco’s modified Eagle’s medium (DMEM) supplemented with 10% heat-inactivated FBS, 1% pen/strep, 2mM L-glut, 1% non-essential amino acids, 1% HEPES. Severe acute respiratory syndrome coronavirus 2 (SARS-CoV-2, Isolate USA-WA1/2020) was acquired from BEI Resources (NR-52281) and propagated in Vero E6 cells for 3 days. Infectious virus was isolated by harvesting cellular supernatant and spinning at 1000 rpm for 10 minutes to remove cellular debris and stored at −80°C. SARS-CoV-2 infectious particles were quantified in media by standard plaque assay.^46^ Briefly, Vero E6 cells were treated with serial dilutions of SARS-CoV-2 in DMEM (4% FBS) and incubated at 37°C for 1 hr. An overlay of 2.4% cellulose (colloidal microcrystalline, Sigma #435244) and 2x DMEM (5% FBS) was added to each well and incubated for 3 days at 37°C and 5% CO2. On day 3, cells were fixed with 4% paraformaldehyde and stained with crystal violet to visualize and count plaques.

### Oligomer and polymer incubations with SARS-CoV-2

Individual compounds were diluted to 10 μg/mL or 5 μg/mL in DMEM culture media (4% FBS). SARS-CoV-2 was added to the media solution containing a compound at a final concentration of 2.6×10^5^ pfu/mL. Virus incubated in media was supplemented with a volume of RNA/DNAse-free water equal to that used for the compound dilutions. Samples were then incubated at room temperature inside a Luzchem light chamber and exposed to either no light, bulbs emitting 420 nm light, or bulbs emitting 300-400 nm light for the specified time intervals (see Supporting Information for spectral distributions and irradiance). The concentration of infectious virus present in the samples was then quantified by plaque assay.

## Data Availability

Data can be provided upon request.

## Acknowledgement

AMK was supported by NIH grant 1K22AI141680-01A1. KSS acknowledges the Welch Foundation for support (Grant No. AX-0045-20110629).

## TOC Graphic

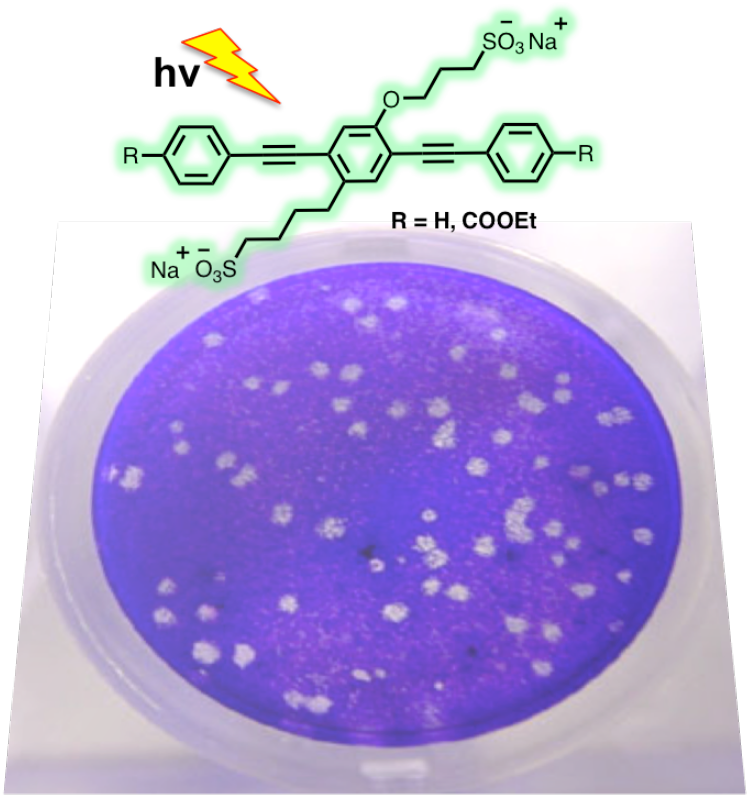

