## Supplemental Information for "Highly Effective Inactivation of SARS-CoV-2 by Conjugated Polymers and Oligomers"

### Materials

All starting materials and reagents were obtained from commercial sources (Sigma-Aldrich, Fisher Scientific) and used without further purification. All reactions were performed under a nitrogen atmosphere, unless stated otherwise. Compounds **1**, **2b, 3**, **poly-4** and **poly-5** were synthesized by following the literature procedure.[1-5]

### Characterization Methods

UV-Visible spectra for all the samples were recorded at a concentration of 10 μg/mL in water using the PerkinElmer Lambda 35 UV-Vis spectrophotometer.

**1,4-bis((4-(3-bromopropoxy)phenyl)ethynyl)benzene (2c).**

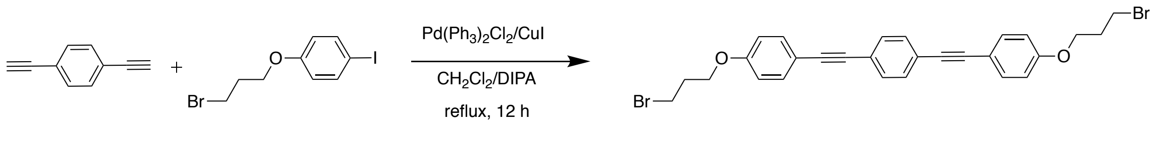


**2a** **2b** **2c**

To a degassed solution of **2a** (300 mg, 2.378 mmol) and **2b** (1.62 g, 4.756 mmol) in 60 mL $CH_2Cl_2$ and 20 mL diisopropyl amine, $Pd(PPh_3)_2Cl_2$ (167 mg, 0.238 mmol), CuI (90 mg, 0.476 mmol) were added. The mixture was refluxed for 12 h. The product was separated between saturated aq.$NH_4Cl$ and $CH_2Cl_2$, and washed with D.I. water. The $CH_2Cl_2$ layer was dried over anhydrous $Na_2SO_4$ and distilled off the solvent under reduced pressure. The crude product was passed through a silica gel column using hexane/DCM as the mobile phase to isolate **2c** as a pale white solid. Yield: 710 mg (76 %).

**3,3'-((((1,4-phenylenebis(ethyne-2,1-diyl))bis(4,1-phenylene))bis(oxy))bis(propane-3,1-diyl))bis(1-methyl-1*H*-imidazol-3-ium) bromide (2).**

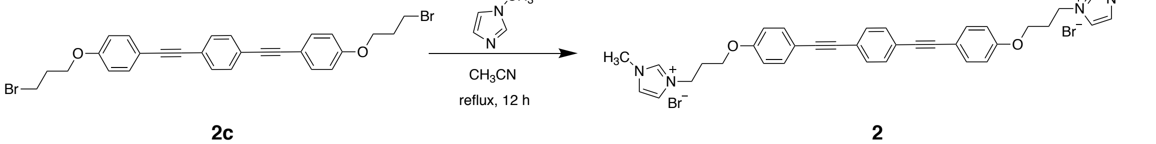


**2c** **2**

To a solution of **2a** (500 mg, 0.905 mmol) in 30 mL $CH_3CN$, 1-methylimidazole (164 mg, 2 mmol, 2.2 eq.) was added and refluxed for 12 h. The precipitated solid product was filtered and was washed with cold $CH_3CN$. The product was dried under high vacuum to remove any trace amount of solvent. Yield: 600 mg (93 %). HRMS (**2**$^{2+}$ ion calculated 278.1413, found 278.1427).

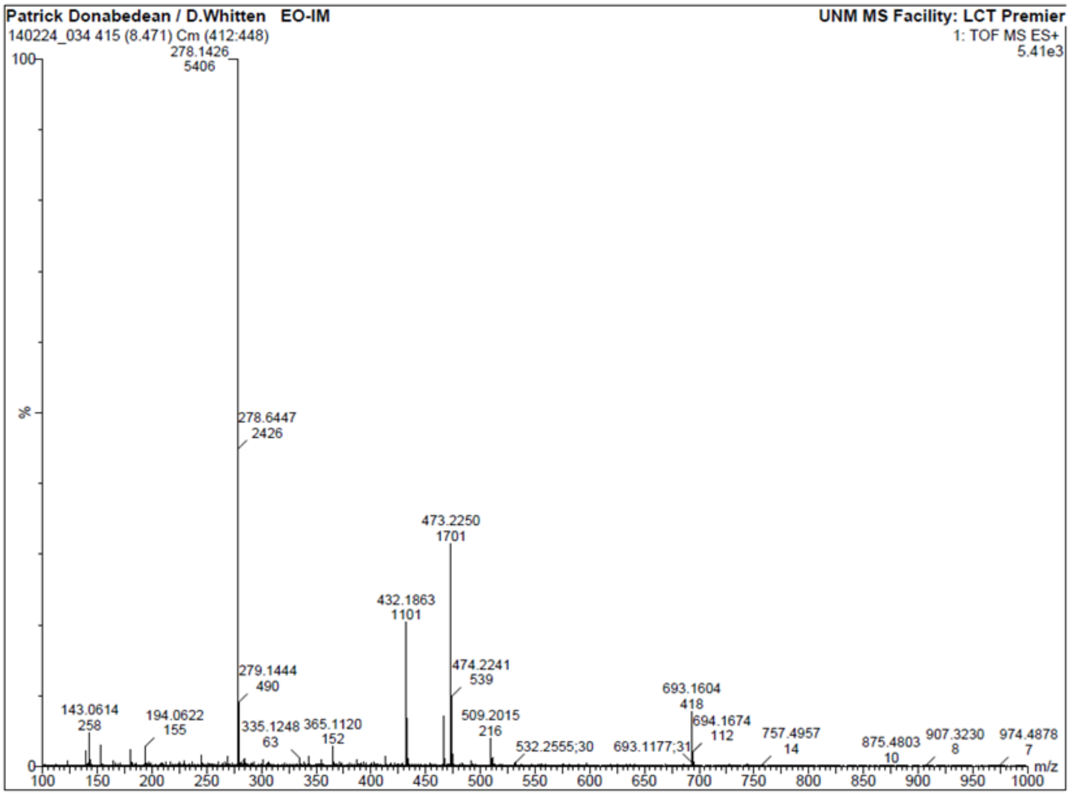


### Photolysis Light Sources

Samples were exposed to light for the indicated periods using using a Luzchem photoreactor (Luzchem.com) equipped with either near-UV or visible light sources. The spectral distribution of the light from the two sources is shown below.

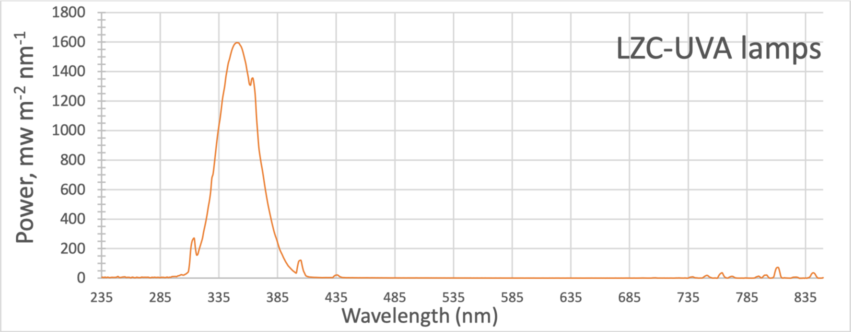


**Figure S-1**. Spectral distribution for LZC-UVA lamps used for near-UV irradiation of samples. The irradiance at the sample with this light source is 6.7 mW/cm$^{-2}$.

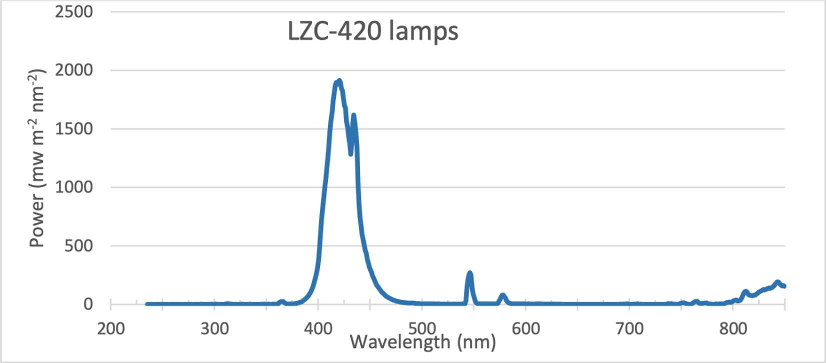


**Figure S-2**. Spectral distribution for LZC-420 lamps used for visible irradiation of samples. The visible region irradiance at the sample with this light source is 6.7 mW/cm$^{-2}$.
